# Clinical Utility of a Highly Sensitive Lateral Flow Immunoassay as determined by Titer Analysis for the Detection of anti-SARS-CoV-2 Antibodies at the Point-of-Care

**DOI:** 10.1101/2020.07.30.20163824

**Authors:** Amanda Haymond, Claudius Mueller, Hannah Steinberg, K. Alex Hodge, Caitlin Lehman, Shih-Chao Lin, Lucia Collini, Heather Branscome, Tuong Vi Nguyen, Sally Rucker, Lauren Panny, Rafaela Flor, Raouf Guirgus, Richard Hoefer, Giovanni Lorenzin, Emanuel Petricoin, Fatah Kashanchi, Kylene Kehn-Hall, Paolo Lanzafame, Lance Liotta, Alessandra Luchini

## Abstract

Coronavirus disease 2019 (COVID-19), caused by the severe acute respiratory syndrome coronavirus-2 (SARS-CoV-2), became a pandemic in early 2020. Lateral flow immunoassays for antibody testing have been viewed as a cheap and rapidly deployable method for determining previous infection with SARS-CoV-2; however, these assays have shown unacceptably low sensitivity. We report on nine lateral flow immunoassays currently available and compare their titer sensitivity in serum to a best-practice enzyme-linked immunosorbent assay (ELISA) and viral neutralization assay. For a small group of PCR-positive, we found two lateral flow immunoassay devices with titer sensitivity roughly equal to the ELISA; these devices were positive for all PCR-positive patients harboring SARS-CoV-2 neutralizing antibodies. One of these devices was deployed in Northern Italy to test its sensitivity and specificity in a real-world clinical setting. Using the device with fingerstick blood on a cohort of 27 hospitalized PCR-positive patients and seven hospitalized controls, ROC curve analysis gave AUC values of 0.7646 for IgG. For comparison, this assay was also tested with saliva from the same patient population and showed reduced discrimination between cases and controls with AUC values of 0.6841 for IgG. Furthermore, during viral neutralization testing, one patient was discovered to harbor autoantibodies to ACE2, with implications for how immune responses are profiled. We show here through a proof-of-concept study that these lateral flow devices can be as analytically sensitive as ELISAs and adopted into hospital protocols; however, additional improvements to these devices remain necessary before their clinical deployment.

## Introduction

In March of 2020, the novel coronavirus disease 2019 (COVID-19) joined the ranks of other infectious diseases such as influenza and plague to be categorized as a pandemic. As of July 13, 2020, roughly 13 million people worldwide have been confirmed to have contracted COVID-19, leading to over 500,000 fatalities so far (1). While vaccine and therapeutic interventions are under investigation by almost every qualified scientific body worldwide, there remain significant challenges associated with reducing spread prior to effective therapeutic strategies. One hurdle to reducing spread of the disease is the difficulty in the identification of both actively infected individuals as well as those who have recovered. Active infections, typically diagnosed by real time reverse transcription polymerase chain reaction (RT-PCR), have been plagued with false negative issues as well as availability issues in some countries, including the United States (2–4). Time since infection has proven to be an important source of variability in real time RT-PCR results due to difficulties in detecting SARS-CoV-2 RNA in patients prior to development of symptoms, or more than 5 days post-development of symptoms (5).

Rather than using real time RT-PCR to constantly test for active cases, serologic antibody testing has been proposed as an important tool to determine the prevalence and spread of SARS-CoV-2 in a community. Positivity rates for the detection of anti-SARS-CoV-2 IgG or IgM in patient serum have been shown to rise above 90% after an average of 11-13 days from symptom onset, though seroconversion timeframes for IgG as it relates to IgM are not necessarily comparable between patients (6). Because antibody testing can be quite variable, there have been concerted efforts to standardize testing. A best-practice ELISA protocol from the Krammer group at Mt. Sinai has been given EUA approval from the FDA, and reagents from this lab have been widely distributed through BEI Resources (7, 8). Additionally, the National Institutes of Health (NIH) has released a broadly similar optimized ELISA protocol that is being adopted for serostudies conducted through the NIH (9). Other groups have reported using similar ELISA protocols with the recombinant HEK293-produced spike receptor binding domain (RBD) protein as the antigen for detection (10). Despite the complications of inter-patient variability and assay timing, ELISA antibody tests have been able to successfully detect robust IgG responses in the vast majority of SARS-CoV-2 infected individuals regardless of clinical severity (6, 7, 11–14). For this reason, serology testing remains a promising lead for understanding the incidence and spread of COVID-19, difficulties notwithstanding.

One approach to overcoming some of the challenges associated with serologic antibody testing is the introduction of lateral flow immunoassays (LFIs): cheap devices that can be rapidly produced and given/interpreted by medical professionals without the need for a laboratory (15). Because of the low cost and high availability, LFIs have the potential to be widely deployed. There have been several studies comparing different commercially-available LFI tests to a best-practice ELISA to determine appropriate candidates for more widespread deployment; one commonly encountered problem is unacceptably low sensitivity (16–18). A recent systematic review and meta-analysis of 40 such studies found that LFIs for COVID-19 had a pooled sensitivity of only 66%, well behind pooled ELISA sensitivity at 84% (19). Moreover, most studies included in the review analyzed LFIs in the laboratory, instead of at the point-of-care (19).

A reason for the overall reduced sensitivity of LFIs versus ELISA assays could be reduced analytical sensitivity of LFIs (20, 21). There are currently no studies directly comparing the analytical sensitivity of multiple LFIs against a best-practice ELISA. Therefore, we proposed to answer two questions: first, are any currently available LFIs as analytically sensitive as an ELISA? Second, if a suitably sensitive test can be identified, is it sufficiently simple to use and to interpret such that it could be deployed clinically? To answer these questions, we report herein on the evaluation of the analytical sensitivity of nine commercially available LFI devices, including two currently under FDA EUA approval, as compared to a best-practice ELISA. Furthermore, we conducted a pilot-scale study of the clinical utility of the most analytically sensitive LFI device at the Santa Chiara Hospital in Trento, Italy and report on the sensitivity and specificity of the device at the point-of-care. This work can be used to inform future studies both on the promise of LFI devices for detection of previous infection with SARS-CoV-2, as well as the hurdles to using these devices clinically.

## Materials and Methods

### George Mason University Patient Cohort

Patients were recruited from the Northern Virginia area and consented under an IRB-approved protocol (Liotta PI, 1592168-1); inclusion criteria for the research study included confirmed or suspected infection with SARS-CoV-2 based on symptoms, contacts, or travel history. A total of seven PCR-positive or clinically-diagnosed patients donated serum samples that were used in this study; four patients donated samples at two different time points. On the day of donation, each patient was consented and donated at least two tubes of blood, one collected in a gel serum separator tube (SST, gold top 5 mL, BD Vacutainer cat# 368013) and one collected in a clot-activator serum tube (RT, red top 5 mL, BD Vacutainer cat# 367814). Blood was allowed to clot for 45-60 minutes at room temperature, and serum was separated via centrifugation (2250 x g, 15 minutes). Serum was aliquoted and stored at -80°C until analysis.

### ELISA

All patient samples were analyzed by ELISA using a protocol modified from (7), with all changes noted below. Patient serum samples were heat inactivated for 30 min at 56°C prior to analysis to ensure samples were non-infectious. ELISA plates (Immulon 1B, ThermoFisher cat# 3355) were coated overnight at 4°C with 2 μg/mL SARS-CoV-2 spike RBD-mFc tag (Sino Biological cat# 40592-V05H) produced in HEK293 cells. Serum samples were incubated on antigen coated plates at 10-fold dilutions ranging from 1:2 to 1:20,000 for 2 hr at room temp. Binding was detected using 1:5,000 goat anti-human IgG-HRP conjugated secondary (Jackson Immunoresearch cat# 109-035-098) with TMB substrate (Fisher cat# 34028) for 12 minutes, with the reaction halted via 2 M sulfuric acid. OD450 values were measured on a plate reader. To correct for intra-plate differences, four calibrator samples consisting of three pre-diluted positive control sera and a negative control were added to every plate. Samples were normalized to the geometric mean of the calibrator values. A total of 12 negative serum samples were used to determine assay background; samples were collected prior to 2019 as healthy controls for an unrelated breast cancer study. Average of the negative samples plus three standard deviations at each dilution was used as the background value. Samples were considered above background if the mean value minus average standard deviation for the sample was greater than the background value at a given serum dilution; differences less than 0.05 abs units were considered insignificant.

### Lateral Flow Immunoassay Titer Analysis

Combined anti-SARS-CoV-2 IgG/IgM lateral flow immunoassays were obtained from the following manufacturers: Pinnacle Biolabs SARS CoV-2 IgG/IgM One Step Rapid Test Device (Pinnacle); Wuhan EasyDiagnosis Biomedicine Co., Ltd COVID-19 (SARS-CoV-2) IgM/IgG Antibody Test Kit, reference SA-2-D (EDiagnostics); SafeCare Bio-Tech COVID-19 IgG/IgM Rapid Test Device (WB/S/P) ref NCO-4022 (SafeCare), AcroBiotech 2019-nCoV IgG/IgM Rapid Test Cassette reference INCP-402 (AcroBiotech); LumiQuick Diagnostics Quick Profile 2019 nCoV IgG/IgM Test Card ref 71108B (LumiQuick); Cellex qSARS-COV-2 IgG/IgM Rapid Test (Cellex); CALTH Care Health AllCheck COVID-19 IgG/IgM reference CHR09E (AllCheck); and Healgen COVID-19 IgG/IgM (Whole Blood/Serum/Plasma) Rapid Test Device reference GCCOV-402a (Healgen). Separate anti-SARS-CoV-2 IgG and IgM tests were obtained from RayBiotech: Coronavirus (SARS-CoV-2) IgG Test Kit ref CG-COV-IgG (RayBiotech IgG), and Coronavirus (SARS-CoV-2) IgM Test Kit ref CG-CoV-IgM-FP (RayBiotech IgM). Tests were conducted as recommended by the manufacturers, with a few modifications. Inactivated serum (20 μL for Pinnacle, EDiagnostics, SafeCare, AcroBiotech, and RayBiotech; 10 μL for Cellex and AllCheck, 5 uL for Healgen, and 2 μL for LumiQuick as recommended in each instruction manual) were pipetted directly on the absorbent sample pad of each test. Immediately following sample deposition, 2-3 drops of buffer included in each kit were added to the sample pad to start the flow of the sample up the lateral flow strip. Results were scored between 10-20 minutes following sample deposition for every test by three independent scorers using a 3-point scale. Absence of a line, or a negative result, was scored as 0; clear presence of a line, or a positive result, was scored as a 2; and a faint line that neared the limit of detection for the scorer or was difficult to see was scored as a 1. All scores were averaged, and the score for a given test was considered positive if the average of the three scores was greater than 1. Titers were determined for each test as the highest dilution of serum that scored greater than 1 for a given patient. Agreement between readers for LFI data was determined via Fleiss’ kappa statistic, Krippendorff’s alpha statistic, and percent agreement as calculated by STATA 13 (StataCorp. 2013. *Stata Statistical Software: Release 13*. College Station, TX: StataCorp LP) for three readers with three variable assignments. For Fleiss’ kappa, 0.4 < κ < 0.75 indicated mild to moderate agreement, and κ > 0.75 indicated strong agreement as recommended in (22).

### Viral Neutralization Assay

Viral neutralization assays were conducted under BSL-3 conditions at George Mason University’s Biomedical Research Laboratory using standard methods. Briefly, SARS-CoV-2 USA-WA1/2020 isolate (BEI Resources, Cat # NR-52281) was added to twofold serially diluted sera at a concentration of 94 pfu/well. Vero cells were infected using the diluted sera and virus mixture and were incubated for 1h at 37°C. Following infection, a 1-mL overlay of a 1:1 solution of 0.6% agarose in diH2O and 2x EMEM supplemented with 5% FBS, 1% L-glutamine, 1% penicillin/streptomycin, 1% non-essential amino acids, and 1% sodium pyruvate was added to the cells. The cells were incubated at 37°C and 5% CO2 for 48 hours. After 48 hours, the cells were fixed using 10% formaldehyde for 1 hour at room temperature. Following fixation, the formaldehyde and agar plugs were discarded. The cell monolayers were stained with a solution of 2% crystal violet and 20% ethanol to visualize plaques. Percent neutralization was calculated as: [(number of SARS-CoV-2 plaques per well without anti-SARS-CoV-2 serum) – (number of SARS-CoV-2 plaques per well of diluted anti-SARS-CoV-2 serum) / (number of SARS-CoV-2 plaques per well without anti-SARS-CoV-2 serum) x 100]. Neutralizing titers were expressed as the reciprocal of the highest dilution of serum that neutralized 90% of SARS-CoV-2. Samples with PRNT90> 1:160 were considered positive, as recommended for convalescent plasma donation by the FDA (23).

### Western Blot Analysis

Serum samples from patients 1F, 8F, 10F, and an uninfected negative control individual were examined for the presence of anti-ACE2 or anti-S antibodies. All samples for this analysis were provided from the first donations of each patient collected in gel serum separator tubes (SST). Whole cell extracts from A549 and Vero E6 were analyzed via BCA assay per the manufacturer’s recommendations (Thermo Fisher Scientific). For western blot, equal amounts of protein were mixed with NuPAGE LDS sample buffer and NuPAGE sample reducing agent, heated at 70°C, and separated on a NuPAGE 4-12% Bis-Tris protein gel (Thermo Fisher Scientific). Proteins were transferred to a Nitrocellulose membrane using the iBlot 2 gel transfer device. Membranes were blocked with 5% milk in TBS with 0.1% Tween 20. Appropriate antibodies or patient samples were added for overnight incubation at 4°C. Membranes were washed, incubated with the appropriate secondary antibodies at 4°C, and developed with either Clarity or Clarity Max Western ECL Substrate (Bio-Rad). Chemiluminescence imaging was performed using either the Azure cSeries imaging system (Azure Biosystems) or the ChemiDoc Touch Imaging System (Bio-Rad). Antibodies used for these experiments included α-Actin (Abcam # ab-49900).

### Clinical Analysis of Lateral Flow Immunoassay

Lateral flow immunoassays from two manufacturers (RayBiotech and SafeCare) were sent to the Santa Chiara Hospital in Trento, Italy for analysis in the clinical setting. PCR-positive patients who had been in the hospital greater than 10 days (n = 27) were consented under IRB compliance. Each patient provided a saliva sample by depositing the sample in a microcentrifuge tube. Saliva samples were centrifuged to separate cellular/food debris from the supernatant. Saliva supernatant was aspirated into the kit-provided plastic droppers to the fill point recommended by the LFI manufacturer and deposited on the absorbent sample pad of the LFI test. Fingerstick blood samples were obtained by piercing the skin with a sterile lancet, and a single drop of blood was expressed and spotted onto the absorbent sample pad of the LFI test. Following deposition of either the saliva or the fingerstick blood sample, 2-3 drops of included sample buffer was added directly to the sample pad to start the flow of sample up the lateral flow strip. Test strips were read at 20 minutes post sample deposition and were scored upon visual inspection by with agreement from at least two nurses. Seven PCR-negative hospitalized patients provided saliva and blood samples as the negative controls. To determine sensitivity and specificity of LFI tests in the healthcare setting for saliva and fingerstick blood, receiver-operator characteristic (ROC) curves were generated with GraphPad Prism version 8.4.3. Two by two tables used to determine positive predictive value (PPV), negative predictive value (NPV), sensitivity, and specificity were generated using Microsoft Excel.

## Results

### Lateral flow immunoassay analysis of PCR-positive patient serum indicates some lateral flow kits can reach ELISA sensitivity levels

A total of five RT-PCR positive patients (1F, 8F, 10F, 20M, and 27F), as well as two patients with a clinical COVID-19 diagnosis (7F and 19F), donated serum towards our analytical sensitivity study comparing a best-practice ELISA assay versus nine commercially available LFI devices. While Patient 7F and Patient 19F were both clinically diagnosed based on symptoms, Patient 19F additionally shared a household with a RT-PCR positive patient 20M. Patients 1F, 7F, 19F, and 20F provided two samples for a total of 11 unique samples for analysis. Table 1 reports the clinical details for each included patient. The first serum donations collected from the first six donors (1F, 7F, 8F, 10F, 19F, 20M) were used to screen all nine lateral flow devices; devices that performed well were brought forward to a secondary screen, which included the remaining five serum samples. We adopted a similar ELISA protocol to that of the Krammer group and the NIH (8, 9) with minimal modifications. Primarily, we conducted a single-tiered screening protocol using the spike receptor binding domain (RBD, Sino Biological) rather than a two-tiered screening protocol using first the RBD, then the full-length spike (S) protein. It has been shown that convalescent patients possessing S-reactive antibodies also possess RBD-reactive antibodies (7).

**Table 1.** Patient Characteristics

| **PATIENT**<br>**#** | **COVID-19 DIAGNOSIS:**<br>**PCR-CONFIRMED,**<br>**CLINICAL, OR NONE** | **SYMPTOMS:**<br>**NONE, WEAK, MODERATE** | **TIME BETWEEN FIRST**<br>**AND SECOND**<br>**DONATIONS** |
| --- | --- | --- | --- |
| **1F** | RT-PCR Confirmed | Moderate | 3 weeks |
| **7F** | Clinical Diagnosis:<br>Based on Symptoms | Mild | 6 weeks |
| **8F** | RT-PCR Confirmed | Moderate | N/A |
| **10F** | RT-PCR Confirmed | Moderate | N/A |
| **19F** | Clinical Diagnosis:<br>Based on symptoms and<br>RT-PCR+ family members | Mild | 3 weeks |
| **20M** | RT-PCR Confirmed | Moderate | 3 weeks |
| **27F** | RT-PCR Confirmed | Moderate | N/A |

As shown in Figure 1, five of the six patients were determined to have anti-SARS-CoV-2 RBD-reactive IgG antibodies at levels above assay background (dotted line in Figure 1) via ELISA. Patient 7F did not have detectable antibodies above background via ELISA at any dilution, and thus could not be included in the analytical sensitivity analysis. ELISA titers detected in patients 1F, 8F, 10F, 19F, and 20M range between 1:200 to 1:20,000 dilution of serum. The lowest dilution of serum which was positive above background via ELISA provided the “benchmark” for the titers for a patient; this benchmark is noted with a gold line in Figure 1. LFIs were then evaluated for their analytical sensitivity and compared to this ELISA benchmark.

**Figure 1.**
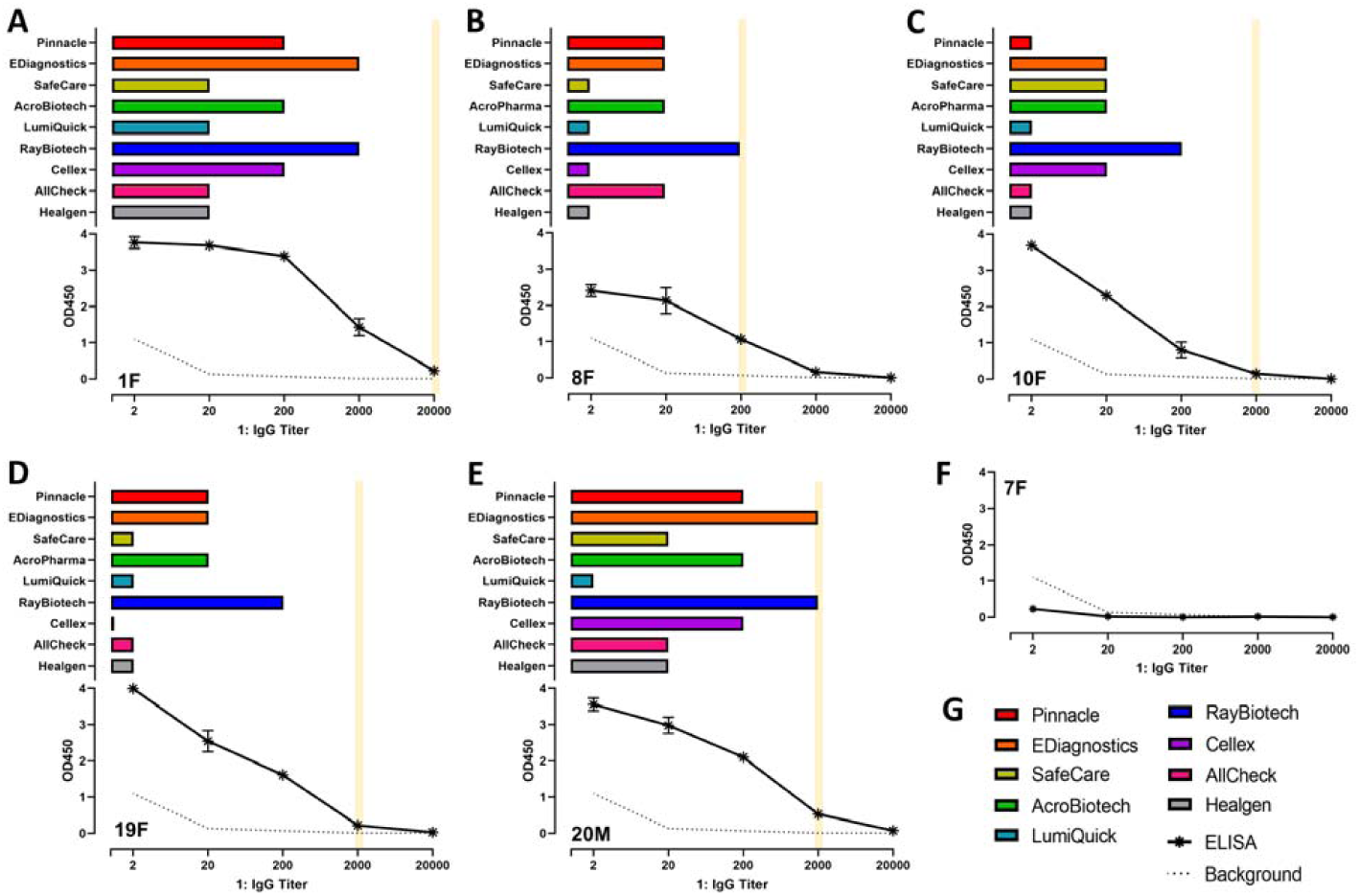
Titer sensitivity of nine LFIs compared to a best practice ELISA reveals two devices with high analytical sensitivity. Negative control samples (N = 12) were used to determine ELISA assay background. A) Patient 1F showed titers down to 1:20,000 above background via ELISA. EDiagnostics LFI and RayBiotech LFI were the most sensitive for this patient, showing positive scores to 1:2000. B) Patient 8F showed titers down to 1:200 above background via ELISA and RayBiotech LFI. C) Patient 10F showed titers down to 1:2000 above background via ELISA. RayBiotech LFI was the most sensitive for this patient, showing positive scores to 1:200. D) Patient 19F showed titers down to 1:2000 above background via ELISA. RayBiotech LFI was the most sensitive for this patient, showing positive scores down to 1:200. E) Patient 20F showed titers down to 1:2000 above background via ELISA, EDiagnostics LFI, and RayBiotech LFI. F) Patient 7F was negative at all dilutions. G) Legend utilized for panels A-F.

For most patients, an IgG LFI device from RayBiotech (Figure 1, in blue) proved the most sensitive of the LFIs tested, hitting the ELISA benchmark shown in gold for two of the five tested patients in the first screen. Additionally, patients 1F (Figure 1A), 10F (Figure 1C), and 19F (Figure 1D) showed values in the ELISA above background only by a very small margin, suggesting that even in patients 8F and 19F, the RayBiotech device was very sensitive. An LFI device from EDiagnostics (Figure 1, in orange) also performed well, with titers equaling that of the ELISA in one of the five patients. Each LFI was tested with dilutions of serum ranging from 1:2 to 1:20,000 as well as undiluted serum. Device scoring was conducted using a 3-point scale in which 0 represented a negative result (absence of a test line), 2 represented a positive result (presence of a test line), and 1 represented an indeterminate result (line at the reader’s visual limit of detection). Three readers independently scored each kit, and the average of the three scores was used to report LFI results (Figure 1 and Supplementary Figure 1). Raw data is reported in Supplementary Table 1. For a particular serum dilution to be considered positive via LFI, the average score from all three readers for that test had to be greater than 1. Because the LFI titer sensitivity is the average of three scores, it should be kept in mind that the titer sensitivity value reported does have intrinsic error, the quantification of which is further discussed in Table 2.

**Table 2:** Concordance between LFI readers. Fleiss’ kappa, Krippendorff’s alpha, and percent agreement values describing reader agreement by test. All samples (both IgM and IgG results) are considered.

| MANUFACTURER | FLEISS'<br>KAPPA | KRIPPENDORFF'S<br>ALPHA | PERCENT<br>AGREEMENT |
| --- | --- | --- | --- |
| PINNACLE | 0.868 | 0.869 | 0.922 |
| EDIAGNOSTICS | 0.500 | 0.501 | 0.691 |
| SAFECARE | 0.747 | 0.748 | 0.880 |
| ACROBIOTECH | 0.801 | 0.802 | 0.900 |
| LUMIQUICK | 0.916 | 0.917 | 0.967 |
| RAYBIOTECH | 0.399 | 0.401 | 0.611 |
| CELLEX | 0.587 | 0.590 | 0.776 |
| ALLCHECK | 0.650 | 0.652 | 0.822 |
| HEALGEN | 0.648 | 0.650 | 0.800 |
| **ALL TESTS** | **0.659** | **0.671** | **0.800** |

IgG responses have been reported to be more reliable via lateral flow than IgM responses. This is attributed to the low affinity of IgM antibodies and the narrow window in which IgM antibody concentrations are high (24). While all IgG LFIs were positive with undiluted serum for all patients, there was a marked reduction in agreement between LFI devices for the presence of anti-SARS-CoV-2 IgM antibodies, even with undiluted serum. As shown in Table S1, some devices showed higher scores at lower dilutions (i.e. 1:20 versus undiluted), complicating analytical sensitivity analysis. The strict time dependence for detection of IgM antibodies and known reduced sensitivity of IgM versus IgG LFIs for the detection of previous SARS-CoV-2 infection (16) led us to focus on IgG results as the more clinically relevant antibody isotype, with IgM results presented in the supplementary Table S1 and Figure S1.

Based on the results of the first screen, a secondary screen with the remaining five samples was conducted with the highest sensitivity LFIs, EDiagnostics and RayBiotech, along with a representative from the lower sensitivity LFI group, SafeCare, which reports using the same antigen as the RayBiotech test. For the secondary screen, one new patient was tested (27F), as well as repeat donations from 1F, 7F, 19F, and 20M, marked as “D2” for second donation. As shown in Figure 2, the EDiagnostics and RayBiotech test kits were again found to be more analytically sensitive than the SafeCare test. Patient 7F again tested negative via all LFIs and ELISA. For LFI+ patients who donated two samples (1F, 19F, 20M), no significant difference in titer level was detected between donation dates, which were three weeks apart for these three patients. The EDiagnostics test was as analytically sensitive as the ELISA in three patients’ samples (1F:D2, 20M:D2, and 27F), while the RayBiotech test was as analytically sensitive as the ELISA in one patient’s samples (1F:D2).

**Figure 2:**
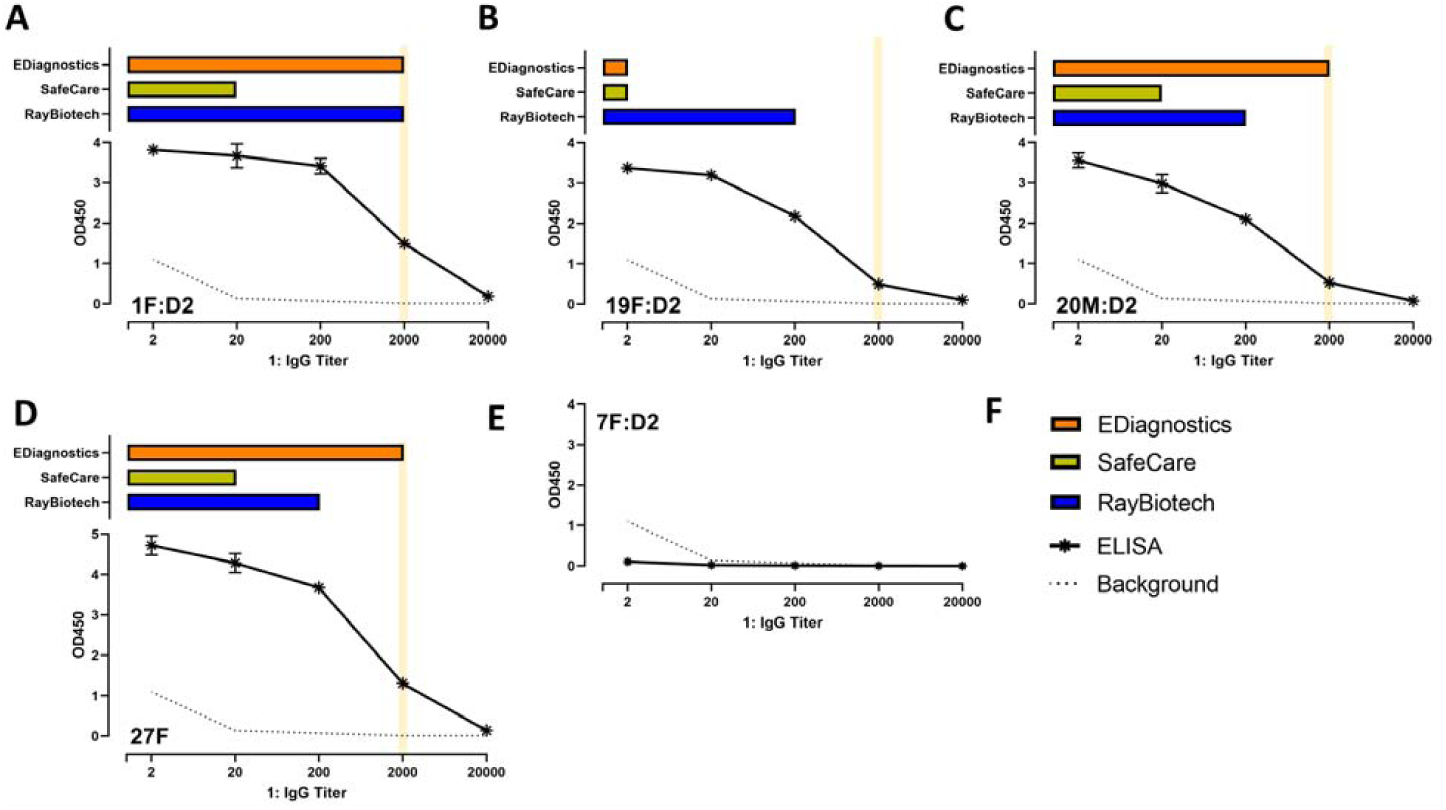
Comparing titer sensitivity of three LFIs, two high performing in the primary screen and one average-performing in the primary screen, reveals continued high analytical sensitivity of LFIs from EDiagnostics and RayBiotech. Negative control samples (N = 12) were used to determine ELISA assay background as in Figure 1. No significant difference in titers between first and second patient’s donations was observed. A) Patient 1F, second donation sample, showed titers down to 1:2000 above background via ELISA, EDiagnostics LFI, and RayBiotech LFI. B) Patient 19F, second donation sample, showed titers down to 1:200 above background via ELISA and RayBiotech LFI. C) Patient 20M, second donation sample, showed titers down to 1:2000 above background via ELISA and EDiagnostics LFI. D) Patient 27F showed titers down to 1:2000 above background via ELISA and EDiagnostics LFI. E) Patient 7F, second donation sample, was negative at all dilutions. F) Legend utilized for panels A-E.

Only four of the nine manufacturers have directly reported the antigens used as bait for the patient IgG and IgM. Both SafeCare and RayBiotech report using solely the N protein, Healgen reports using the S protein, and Cellex reports using both N and S. This is of particular interest as the RayBiotech test showed roughly equivalent titer sensitivity to the in-house ELISA, despite using a different antigen (N vs S RBD). Previous reports comparing SARS-CoV-2 antibody ELISAs utilizing S RBD protein versus N protein have shown increased sensitivity and specificity when using the S protein (10).

When comparing the nine tests, it becomes clear that there can be a trade-off between sensitivity and line intensity. Table 2 reports the Fleiss’ kappa, Krippendorff’s alpha, and percent agreement values for each test, including combined results for both IgG and IgM for all LFIs tested in the first and second screens (IgG and IgM values are reported separately in Table S2 and S3). These metrics describe the relative agreement in scores given by each of the three readers. Focusing on the kappa value as an example, all tests kits had kappa values greater than 0.4, outside of the RayBiotech test which scored a 0.399, which indicates mild to moderate agreement. Three tests had kappa values greater than 0.75, indicated strong agreement as recommended by (22). While test kits from RayBiotech and EDiagnostics were the most sensitive kits tested (Figure 1), they also displayed the lowest Fleiss’ kappa values (Table 2). Conversely, the test kit with the lowest sensitivity, available from LumiQuick, displayed the highest Fleiss’ kappa value. We can report qualitatively that those test kits with higher Fleiss kappa scores, such as LumiQuick, Pinnacle, and AcroBiotech, had test lines that were more intense and more easily distinguished from background over kits with higher kappa values, leading to improved agreement in scoring. We propose that while the RayBiotech and EDiagnostics kits were the most analytically sensitive, much of that sensitivity is garnered by weak positive results at high dilution, which were inconsistently scored between readers. It is worth considering that while sensitivity has been reported as a key weakness of lateral flow immunoassay devices, simple increases in sensitivity may not directly translate to clinical performance due to issues in consistent scoring between readers.

### Viral neutralization confirms protection in all ELISA-positive patients, but some infected patients may develop anti-host versus anti-viral antibodies, complicating interpretation

Based on the results of the ELISA titer analysis and LFI titer analysis on a small group of five PCR-positive patients and two clinically-diagnosed patients, we determined that one clinically-diagnosed patient 7F did not possess significant anti-S RBD antibodies (ELISA) or anti-N antibodies (RayBiotech, SafeCare LFIs) in serum above background. Current preprint literature suggests that some patients who are infected produce robust memory T-cell responses even in the absence of seroconversion (25) indicating that lack of an ELISA or LFI response in patient 7F does not indicate lack of SARS-CoV-2 infection. Given the absence of PCR testing in this patient, it is unknown if the patient was truly infected with SARS-CoV-2. To determine if patient 7F mounted an antibody response but failed to make significant anti-S or anti-N antibodies within ELISA or LFI detection limits, we sought additional information by utilizing plaque reduction neutralization tests (PRNT). All other patients were included in the analysis for comparison.

As shown in Figure 3A, all patients with detectable antibodies via ELISA and LFI demonstrated viral neutralization at levels considered suitable for convalescent plasma donation (PRNT90 > 160) in both gel serum separator tubes (SST) and clot activator serum tubes (RT) (23). Patient 7F however had no neutralizing antibodies detected in either sample. Given this patient’s clinical diagnosis based on distinctive symptoms (loss of taste and smell), we decided to probe 7F’s sample for presence for additional factors that indicates infection. There have been reports that some viral neutralization assays conducted with serum from patients in the context of SARS showed interference due to autoantibodies (26), but there have not been reports of this phenomenon yet with SARS-CoV-2 patients. Given this possibility, we conducted Western blot analysis with the serum of patient 7F compared to PCR-positive 1F, 8F, and 10F to determine if patient 7F could be making autoantibodies against receptors for SARS-CoV-2, which could be protective but may not be neutralizing based on the conditions of the PRNT assay. We specifically considered angiotensin converting enzyme-2 (ACE2), the most well-known receptor for SARS-CoV-2 (27). As shown in Figure 3B, patient 7F’s serum reacted strongly with the ACE2+ cellular lysates, with significantly higher intensity observed for ACE2 bands with this patient than any other patient examined. Patient 1F and 8F had lower reactivity to ACE2, while 10F had no detectable reactivity to ACE2. A negative patient without SARS-COV-2 infection also showed no reactivity to ACE2. Conversely, patients 1F and 8F showed anti-S antibodies, confirming the ELISA results from Figure 1, while patient 7F was again negative for S antibodies, as shown in Figure 3C. Interestingly, patient 10F did not have detectable anti-S antibodies in this assay. Based on this data, where known positive patients mounted limited anti-ACE2 antibodies (1F and 8F), we conclude that some patients may produce anti-host antibodies as well to protect against disease. It is possible this was the primary response for patient 7F, explaining why other measures of response were negative. We conclude that if patient 7F was truly infected likely with SARS-CoV-2, as distinctive symptoms would suggest, they did not mount significant antibody response against the virus itself that we could detect, but potentially produced some anti-host antibodies instead. These anti-host antibodies did not appear neutralizing in this assay. Because of the uncertainty of infection in patients who did not receive a positive RT-PCR test but only a clinical diagnosis, we did not include any clinically diagnosed patients as potential positive cases for COVID-19 disease for our point-of-care analysis.

**Figure 3.**
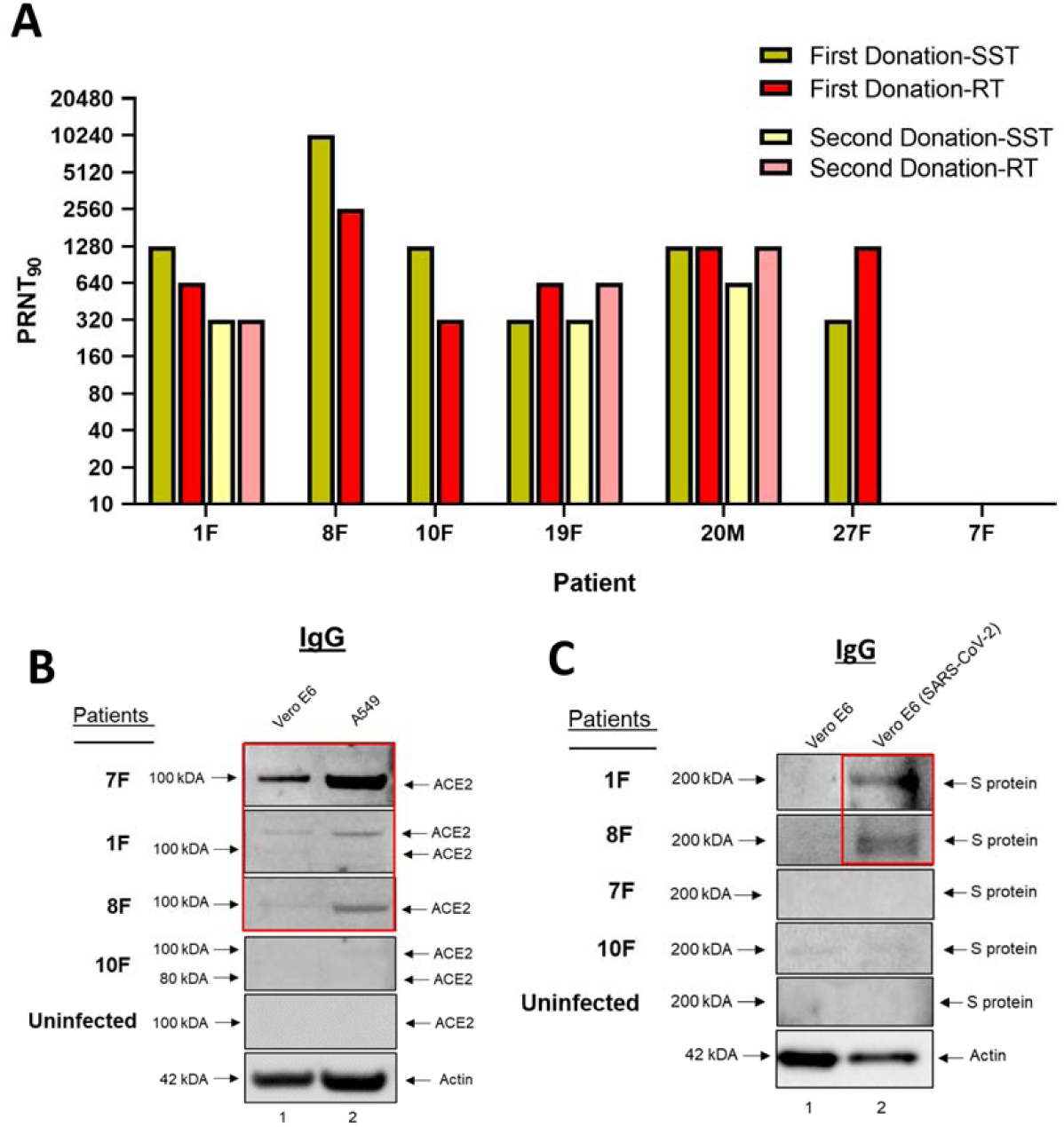
Viral neutralization, ELISA, and LFI negative patients may still harbor anti-receptor antibodies that could affect clinical disease. A) PRNT90 values of PCR-confirmed or clinically diagnosed COVID-19 patients shows high neutralizing titers only for patients who also harbor anti-S or anti-N antibodies as determined via ELISA or LFI. Patient 7F did not inhibit the virus. Both gel serum separator tubes (SST) and clot activator serum tubes (RT) showed broadly similar titers for most patients. B) Patient 7F’s first donation serum sample reacts strongly with ACE2+ lysates, above both other infected and uninfected patients, suggesting the presence of significant autoantibodies against ACE2. C) Patients 1F and 8F’s first donation samples show reactivity against the S protein from infected Vero cells, while 7F and the uninfected control do not. While 10F’s first donation sample showed anti-S responses via ELISA, 10F does not show anti-S antibodies in this assay.

### Highly sensitive lateral flow immunoassay devices used at the point-of-care can distinguish cases from controls but require additional improvements before widespread clinical deployment

Analytical sensitivity of the RayBiotech and EDiagnostics LFI tests were shown to be similar to that of the best-practice ELISA in a total of six patients. To determine if this sensitivity could translate to clinical utility, we tested the RayBiotech LFI devices on 27 PCR-confirmed, hospitalized patients in Trento, Italy. Due to the results reported in Figure 3, only PCR-confirmed patients were considered as true positive cases for this clinical study. Each patient was tested with three lateral flow devices: RayBiotech’s IgG test kit, RayBiotech IgM test kit, and SafeCare’s IgG/IgM test kit as a comparison to a less sensitive test. RayBiotech and SafeCare’s tests both use the N protein for detection, allowing for direct comparison between the two LFI devices. Each patient was tested using fingerstick blood, which both manufacturers recommend, as well as saliva, which is expected to be lower sensitivity and which no manufacturer recommended. As shown in Figure 4B, the IgG LFI from RayBiotech was successfully used in the clinic and could distinguish between cases (N=27) and controls (N=7) better than chance using fingerstick blood. All 2 by 2 tables are reported in Supplementary Figure 2. As expected, saliva was a poorer discriminator between cases and controls than was fingerstick blood using the RayBiotech test; however, it was a better discriminator using the SafeCare test. This improved AUC value in saliva was driven by the 100% specificity, determined by no false positive detection in the small sample (N=7) of negative saliva. Thus, it remains to be seen if this test could be used in saliva clinically. The corresponding sensitivity was particularly poor (48%). Larger studies of saliva will be needed to determine if this biofluid is a promising source of anti-SARS-CoV-2 antibodies for serostudy detection. Based on this pilot scale point-of-care study, the RayBiotech test was superior to the SafeCare test in the accuracy of anti-SARS-CoV-2 IgG detection using fingerstick blood.

**Figure 4.**
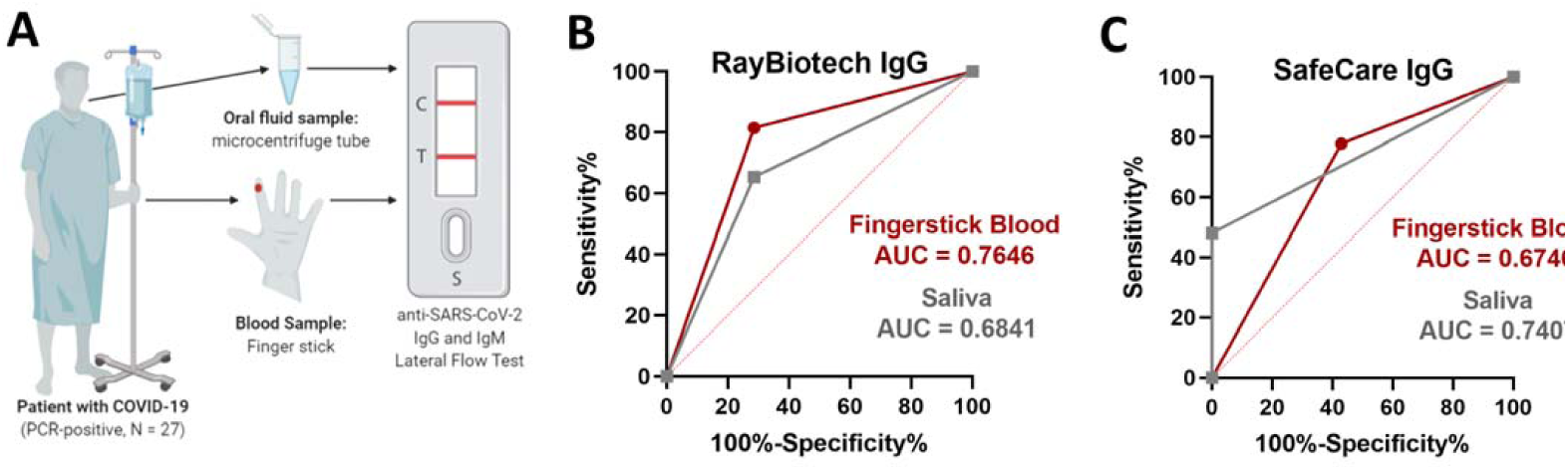
PCR-positive COVID-19 patients tested in the hospital at the point-of-care were correctly identified as positive more often by the RayBiotech test kit than by the SafeCare test kit using fingerstick blood. A) Schematic of trial design. N=27 hospitalized COVID-19 patients greater than 10 days after start of symptoms were consented and donated a fingerstick blood sample and an oral fluid sample. Each patient’s sample was tested against both the RayBiotech IgG and IgM test kits, and the SafeCare IgG/IgM test kit. B) RayBiotech correctly distinguished cases from controls (N=7 hospitalized, negative patients) with an AUC value of 0.7646 for fingerstick blood. Fingerstick blood was superior to saliva as hypothesized. C) SafeCare correctly distinguished cases from controls with an AUC of 0.6746 for fingerstick blood. Saliva was surprisingly superior to fingerstick blood, driven largely by the 100% specificity value. Sensitivity with saliva was poor (48%).

## Discussion

We set out to answer two primary questions in our investigation: first, are there LFI tests that are as analytically sensitive as the best practice ELISA? Secondly, if such a test were to be found, could it show improved point-of-care performance over other reported LFI tests? To answer the first question, we first examined a group of patients (N=7) from the community that were either PCR-positive or clinically diagnosed to investigate the analytical sensitivity of nine different LFI devices, including two devices with FDA EUA approval (Cellex and Healgen), using an ELISA assay as a comparator. To our knowledge, no one has reported a screen of the analytical sensitivity of multiple LFI devices, and several of the LFI devices in our selection, including both the RayBiotech and EDiagnostics LFIs, have not been reported in other publications. We found that the two most analytically sensitive LFIs for this cohort, from RayBiotech and EDiagnostics, showed similar analytical sensitivity to the ELISA. This potentially justifies their use in the clinic, which until now has reported poor sensitivity with LFI devices.

Based on the analytical sensitivity analysis, it may be useful to consider the role these tests could have in screening convalescent plasma for donation. In this scenario, specificity concerns are minimized by only testing PCR-positive patients, while sensitivity concerns are amplified as one wants to identify all potential convalescent plasma donors. Our small patient group utilized in the analytical sensitivity study consisted of exclusively non-hospitalized patients. It may be worth considering using these lateral flow devices as additional tools for identifying potential convalescent plasma donors, as this is much less time intensive and expensive than performing viral neutralization assays on potential donors instead.

To evaluate how much increased sensitivity could improve the accuracy of LFI tests at the point-of-care, we performed a pilot-scale clinical evaluation of the sensitive RayBiotech test against the less sensitive SafeCare test. Both tests report using the N protein as the capture antigen; this ensures an appropriate comparison between the tests. We found that when examining fingerstick blood, the RayBiotech test was more sensitive than the SafeCare test, as hypothesized based on the sensitivity observed in our analytical sensitivity evaluation, but not by as great a margin as expected (81% versus 78%, respectively, Figure S2). Instead, the difference in AUC values was driven more by the increased specificity of the RayBiotech test (71% versus 57%, respectively). Furthermore, it is unclear how significantly the results of such a screen may be complicated by irreproducible scoring; we found that both the EDiagnostics test and the RayBiotech test showed only mild correlation when scored by multiple independent readers in the laboratory. Going forward, the correct balance between sensitivity and consistent readability will be critical for useful clinical adoption.

Both tests, when used on patient saliva, showed reductions in sensitivity as compared to fingerstick blood, as would be expected. While the SafeCare test showed an improved AUC with saliva over fingerstick blood, this was driven by the 100% specificity value on a very small cohort of negative patients (N=7), and it is unclear if saliva would be a suitable biofluid for analysis with this LFI. Larger trials are ongoing to determine if saliva, either raw or processed to concentrate antibodies, can be substituted for serum when using lateral flow devices. Given the noted sensitivity problems even with serum for lateral flow devices, and the fact that saliva contains a greatly reduced concentration of antibodies as compared to serum (28), it is likely that some additional processing steps will be required. There have been reports on the use of saliva for detection of anti-SARS-CoV-2 antibodies (29); to our knowledge, there have not yet been any reports adapting saliva for use in the context of LFI devices for antibody detection. This could represent a large improvement in patient acceptability and ease-of-use for serostudies, however, as we show in our point-of-care analysis, saliva remains unsuitable for this use without further improvements in methodology or devices.

In a surprising turn, during the process of evaluating the viral neutralization potential of patient serum in the analytical sensitivity portion of the study, we did find a noteworthy presence of anti-ACE2 autoantibodies in the serum of patient 7F, as well as limited anti-ACE2 antibodies in patient 1F and 8F’s samples as well. Patient 7F, with the highest concentration of anti-ACE2 antibodies, reported having hypothyroidism, an autoimmune condition. We speculate that autoimmune conditions, such as hypothyroidism, may occur in patients with “overactive” immune systems that are subsequently more likely to harbor autoantibodies such as those detected against ACE2 over other healthy patients such as 1F and 8F. Further research is needed to determine if hypothyroidism, or other autoimmune conditions, could impact the antibody repertoire produced by the host in response to SARS-CoV-2 infection. Furthermore, our rationale for testing against cellular proteins such as ACE2 was that there were either low to no antibodies against S antigen and that viral infections usually cause either partial cell damage or complete cell death (i.e., apoptosis, pyroptosis, etc), where cellular antigens that are normally hidden/protected from the immune system would now become a source of viable antigens for the immune system to recognize (i.e., autoantigens). This is also in line with recognition of the conserved molecular structures expressed by invading microbes named pathogen-associated molecular patterns (PAMPs), and the uncontrolled inflammation in chronic diseases which leads to tissue damage generating endogenous danger-associated molecular patterns (DAMPs) including stress signals such as damaged or apoptotic cells recognized by PRRs (30). Although these signals promote cellular immunity in acute settings (i.e., NLR, RLR, and TLRs), we believe that they could also pose as potential source of antigens in chronic cases of diseases such as COVID-19, where survivors are battling long-term effects (∼20% of all patients). In this case, there are many reports of prolonged symptoms, from chest pain and gastrointestinal issues to cognitive problems and debilitating fatigue that resemble autoantibody related pathologies including Kawasaki disease (KD), the leading cause of acquired heart disease in children; rheumatoid arthritis; inflammatory diseases; and a host of other pathologies including encephalitic syndrome and anosmia (31–34). Future experiments will better define the role of cellular DAMPs (including other receptor/co-receptors such as TMPRSS2, Cathapsin L, CD147, and Neuropilin-1) in COVID-19 infections and whether a robust and aggressive immune response present in these patients is against the pathogen only, or a mixture of DAMPs (i.e., ACE2 antibodies that also block viral entry into cells) that promote immunogenicity and efficacy by enhancing innate and cellular immune responses.

In summary, the poor sensitivity of LFIs in the context of SARS-CoV-2 antibody diagnostics has been documented. Here we demonstrate that this is a resolvable challenge, as several manufacturers have produced very analytically sensitive tests. However, the transition from the laboratory to the clinic reveals additional problems in specificity and ease of use that mandate addressing. Furthermore, as others have reported, not all patients make significant anti-S or anti-N antibodies (35), but may have alternative immune responses to SARS-CoV-2 that are protective. When using antibody tests, one must consider how may truly infected patients could be missed in the analysis due to alternative immune responses, and one may want to consider a suite of multiple viral and host proteins to truly understand a patient’s antibody response. We look forward to additional improvements in these antibody tests to allow them to meet the diagnostic needs of the present moment.

## Data Availability

All data, if not included in supplemental materials, is freely available upon request.

## Acknowledgements

We gratefully acknowledge Dr. Aarthi Narayanan for confirming the reported heat sensitivity of SARS-CoV-2. We thank Dr. Ali Andalibi for his advocacy for our project and support at George Mason’s College of Science. Furthermore, we profusely thank the patients who donated their serum and their time to answer critical questions for the health of others.

## Author Contributions

AH: Performed LFI experiments, analyzed data, wrote paper

CM: Performed the ELISA experiments, analyzed data

HS: Performed the ELISA experiments, analyzed data

AR: Performed the phlebotomy experiments; George Mason University study coordinator

CL: Performed the viral neutralization experiments

SCL: Performed viral neutralization experiments

LC: Performed the clinical point-of-care experiments HB: Performed the western blots

TVN: Performed the ELISA experiments SR: Performed the ELISA experiments

LP: Performed viral neutralization experiments

RF: Performed viral neutralization experiments

RG: Provided reagents (lateral flow test kits)

RH: Provided reagents (control serum samples)

GL: Designed the clinical point-of-care experiments

EP: Designed the patient cohort study, recruited patients

FK: Designed the western blot study, performed the western blots

KKH: Designed the viral neutralization study

PL: Italy study coordinator

LL: Designed the lateral flow immunoassay study, provided funding

AL: Designed the clinical study, analyzed data, provided funding

## Supplementary Figures

**Supplementary Table 1:** Raw patient LFI score data. (Excel sheet of raw data will be uploaded upon submission)

**Supplementary Figure 1.**
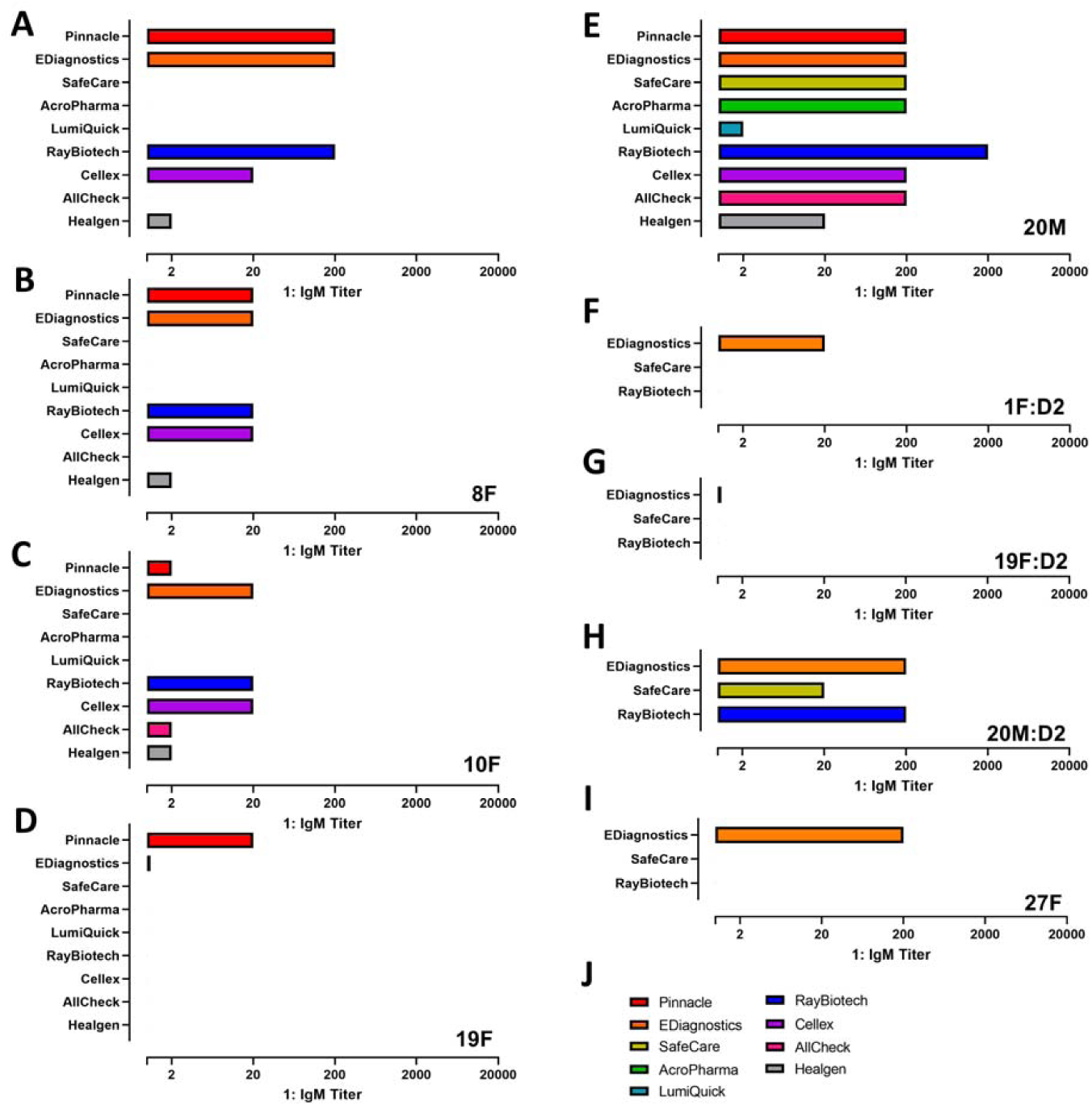
Analytical sensitivity analysis of lateral flow immunoassay devices for IgM with 6 PCR-positive/ELISA-positive/viral neutralization positive patients, including second donations from patients 1F, 19F, and 20M, reveals reduced IgM titers from first donation as expected. A) Patient 1F showed positive results out to 1:200 dilution with three LFIs. B) Patient 8F showed positive results out to 1:20 dilution with four LFIs. C) Patient 10F showed positive results out to 1:20 dilution with three LFIs. D) Patient 19F showed positive results out to 1:20 dilution with one LFI. E) Patient 20M showed positive results out to 1:2000 dilution with one LFI. F) Patient 1F, second draw, showed positive results out to 1:20 with one LFI. G) Patient 19F, second draw, was only positive for IgM with neat serum using one LFI. H) Patient 20M, second draw, showed positive results out to 1:200 with two LFIs. I) Patient 27F showed positive results out to 1:200 with one LFI. J) Legend for panels A-I

**Supplementary Table 2.** Concordance between LFI readers. Fleiss’ kappa, Krippendorff’s alpha, and percent agreement values describing reader agreement by test. IgG only readings are included.

| MANUFACTURER | FLEISS' KAPPA | KRIPPENDORFF'S ALPHA | PERCENT AGREEMENT |
| --- | --- | --- | --- |
| PINNACLE | 0.886 | 0.888 | 0.933 |
| EDIAGNOSTICS | 0.428 | 0.431 | 0.679 |
| SAFECARE | 0.776 | 0.777 | 0.870 |
| ACROBIOTECH | 0.799 | 0.802 | 0.889 |
| LUMIQUICK | 0.954 | 0.954 | 0.978 |
| RAYBIOTECH | 0.278 | 0.282 | 0.586 |
| CELLEX | 0.697 | 0.905 | 0.821 |
| ALLCHECK | 0.873 | 0.874 | 0.933 |
| HEALGEN | 0.741 | 0.744 | 0.856 |
| **ALL TESTS** | 0.683 | 0.692 | 0.812 |

**Supplementary Table 3.** Concordance between LFI readers. Fleiss’ kappa, Krippendorff’s alpha, and percent agreement values describing reader agreement by test. IgM only readings are included.

| MANUFACTURER | FLEISS' KAPPA | KRIPPENDORFF'S ALPHA | PERCENT AGREEMENT |
| --- | --- | --- | --- |
| PINNACLE | 0.850 | 0.852 | 0.911 |
| EDIAGNOSTICS | 0.533 | 0.535 | 0.704 |
| SAFECARE | 0.560 | 0.563 | 0.889 |
| ACROBIOTECH | 0.674 | 0.678 | 0.911 |
| LUMIQUICK | 0.815 | 0.817 | 0.956 |
| RAYBIOTECH | 0.376 | 0.380 | 0.636 |
| CELLEX | 0.3942 | 0.402 | 0.731 |
| ALLCHECK | 0.323 | 0.330 | 0.711 |
| HEALGEN | 0.553 | 0.558 | 0.744 |
| **ALL TESTS** | 0.600 | 0.6003 | 0.798 |

**Supplementary Figure 2.**
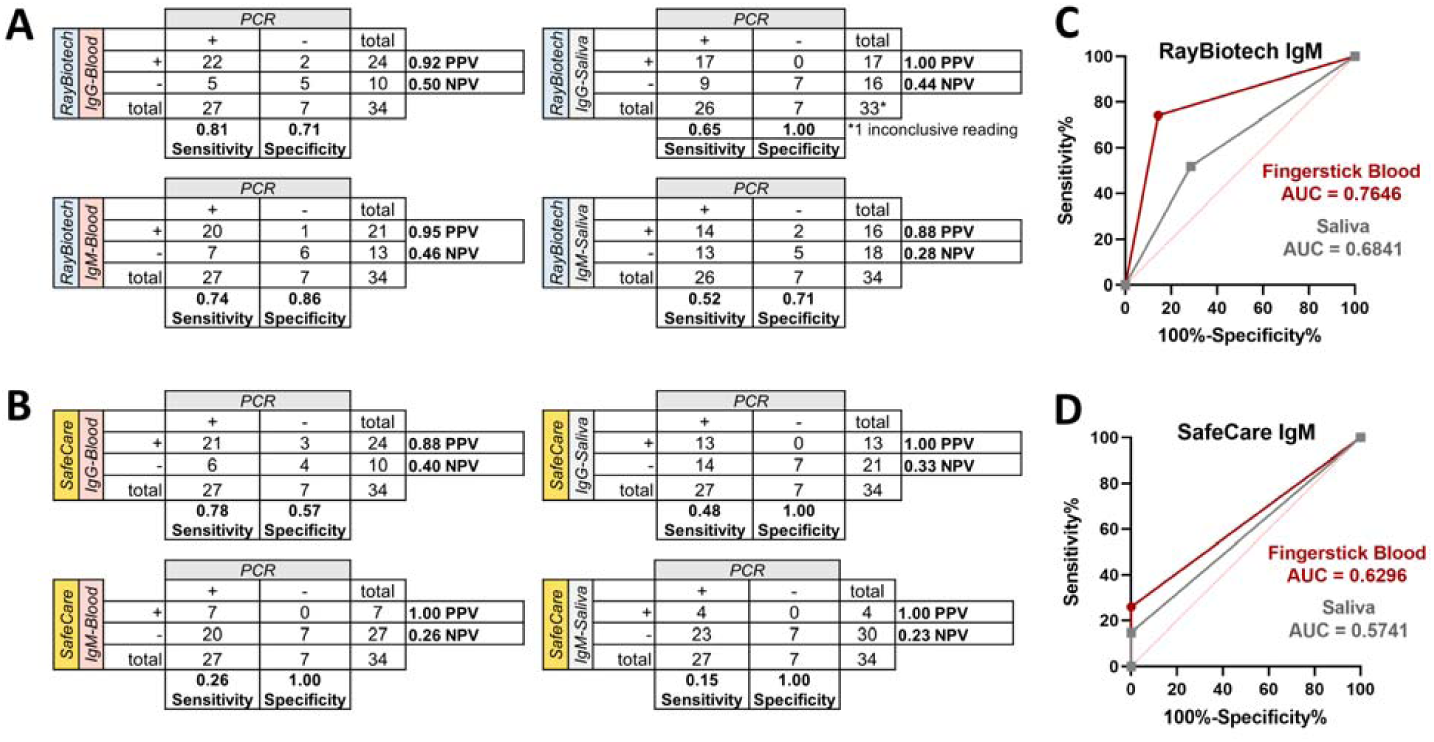
Sensitivity, specificity, PPV and NPV for clinical analysis of RayBiotech and SafeCAre LFI devices. A) 2 × 2 tables for RayBiotech IgG and IgM lateral flow devices when tested on fingerstick blood and saliva. B) 2 × 2 tables for SafeCare IgG and IgM lateral flow devices when tested on fingerstick blood and saliva. C) ROC curve analysis for RayBiotech IgM device. D) ROC curve analysis for SafeCare IgM device.

